# Characterizing the impact of incorporating spatially aggregated human mobility data into infectious disease models

**DOI:** 10.1101/2025.11.26.25340802

**Authors:** Ronan Corgel, Kyra H. Grantz, Lauren Gardner, Derek A. T. Cummings, Harendra de Silva, Thilini Somaratne, Dhammika Silva, LakKumar Fernando, Amy Wesolowski

## Abstract

Models of infectious disease dynamics should align the spatial scale of mobility data to the scale of travel relevant to infer disease introduction events and subsequent local transmission.

Despite this, the biases of spatially aggregating mobility data, which are more commonly available, on model inferences are rarely explored. Here, we examine the sensitivity of infectious disease modeling results to different spatial scales of human mobility by integrating multiscale mobility data from Sri Lanka into SEIR metapopulation models. Aggregated mobility data were obtained from mobile phone records at three increasingly coarser spatial scales to simulate epidemic spread. We found that travel was not evenly distributed amongst nested spatial units when data were disaggregated, with rural subunits exhibiting more external travel than urban subunits. In simulations of non-specific disease transmission, these different scales of mobility aggregation yielded wide variations in estimates of epidemic size and spatial invasion timing. However, modeled differences depended on disease characteristics such as transmissibility and exogenous factors like seeding location urbanicity. Our results carry implications for infectious disease modeling best practices and public health response, particularly the policy decisions made from model inferences that were or were not informed by the relevant spatial scale of mobility data such as intervention timing, risk communication, and resource allocation.

**Significance Statement:** Infectious disease models serve an important role in disease forecasting and outbreak response. As data on human mobility have become increasingly detailed and widely used, the question of spatial scale is paramount to effectively approximating disease dynamics. Our work finds significant discrepancies in estimations of spatial invasion timing and epidemic magnitude simply by changing the scale of mobility data integrated into a transmission model. This could have consequences for estimation of key parameters obtained from such models, the interpretation and communication of infection risk, and ultimately the public health response to a disease threat. We also highlight situations where large model discrepancies are unlikely, such as for highly infectious pathogens and where disease is initially seeded into an urban location.

## Introduction

Human travel by infected individuals has potential to spread infectious diseases, as the COVID-19 pandemic demonstrated.^1,2^ Beyond COVID-19, evidence suggests that spatial disease dynamics for pathogens such as influenza^3,4^, malaria^5^, dengue^6^, yellow fever^7^, cholera^8^, Ebola^9^, and measles^10^ have been driven by the movement of people from one location to another.

Increasingly, data on human movement have been made available from a wide range of sources including mobile phone operators and social media companies.^11^ These data can quantify population mobility at fine temporal resolutions and with wide geographic coverage, even in previously data-sparse regions.^12,13^ With improved access to mobility data, there are more opportunities to uncover the multiscale relationship between human movement and infectious disease transmission (**Figure 1**).

**Figure 1.**
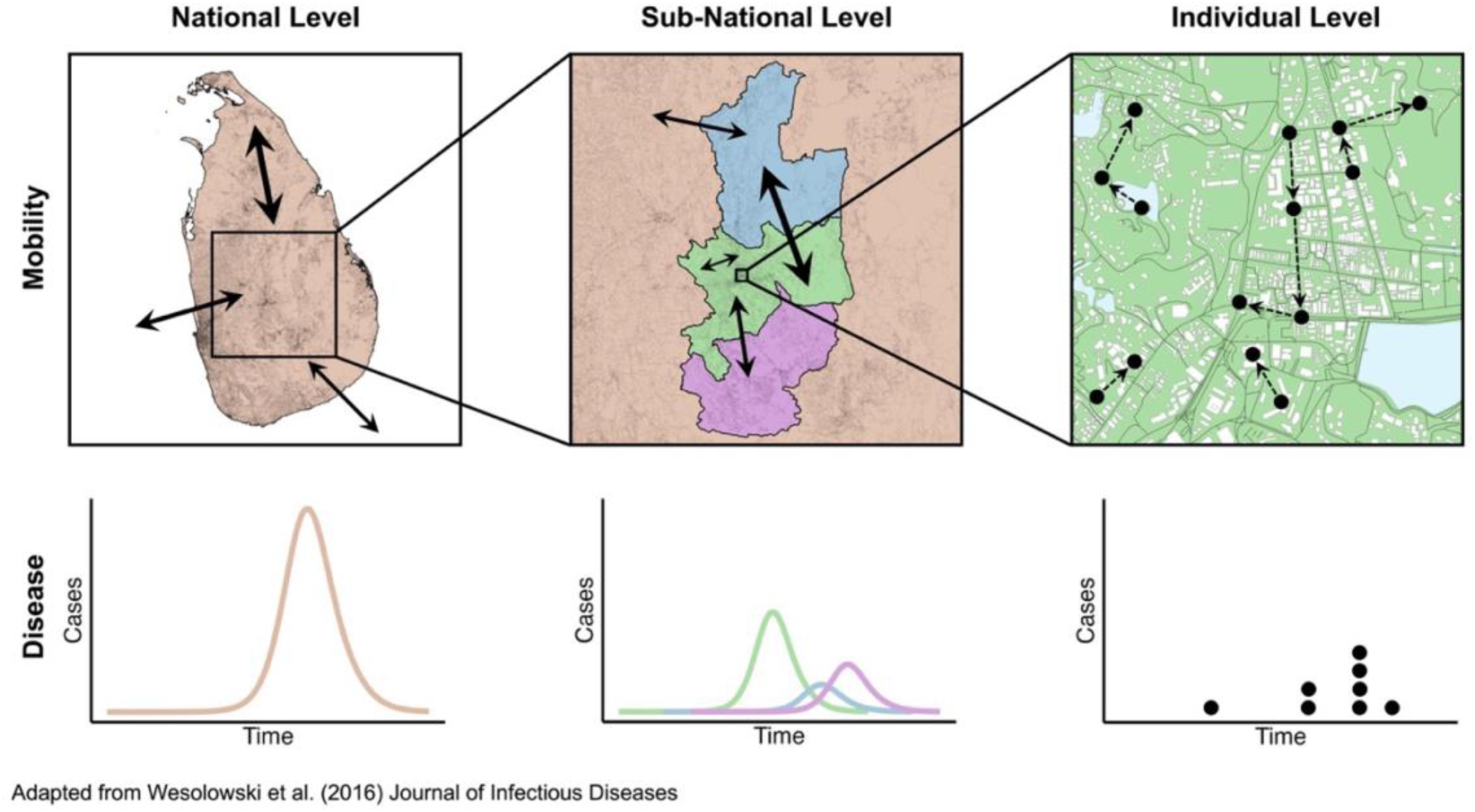
Multiscale mobility and disease data at national, sub-national, and individual levels The schematic depicts the concept that both mobility and disease data can be aggregated or disaggregated to varying spatial scales. Going from left to right, data become increasingly disaggregated to ultimately reach the individual level. Starting at the national level, mobility data describe flows of people leaving, entering, and remaining in a country while disease data depict an epidemic curve of all incident cases recorded for a country. Sub-national mobility data contain information on people leaving, entering, and remaining in units (states, counties, districts, etc.) of a country where more detail is given to within country movement as the number of units increases and the area of units decreases. Similarly, sub-national disease data show the timing and magnitude of epidemic curves for each unit of a country. For the most disaggregated level of mobility data, individual trajectories of people moving throughout space and time are recorded. Individual level disease data include information on each case with potential for added details such as demographics or risk factors. This figure was adapted from a similar figure in a previous work.^14^

Mobility data are often aggregated to quantify travel between administrative units or gridded tiles, and as a function of their granularity, allow for further aggregation to coarser geographic scales.^14,15^ Oftentimes mobility data are available aggregated to a specified spatial unit and as a result, assumptions are made that these trips are evenly distributed amongst smaller, nested spatial units. Data limitations mean that fine scale patterns of importation, exportation, and infection risk may be masked by unit aggregation. Consequently, a specific spatial scale of mobility data, dictated by availability, may inadequately describe disease dynamics and lead to incorrect estimates of disease spatial diffusion and epidemic size. Such misspecifications could in turn complicate public health response efforts, resulting in erroneous outbreak forecasting, communication strategies, and policy interventions.

There is likely an optimal scale of mobility that reproduces infection dynamics. However, finding a scale of mobility detail that sufficiently captures human behavior relevant to disease transmission is complex, since coarser spatial scales may ignore key heterogeneities of travel within units, or conversely, hyper-detailed mobility information may capture individual behavior unassociated with the group patterns that propel disease transmission.^14,17^ For example, recent research found that mobility information at the county scale best approximated the spatial diffusion of COVID-19 in the United States compared to mobility data at increased aggregations.^16^ Additionally, the ideal scale of analysis to infer disease spread will also depend on pathogen epidemiology and the population connectivity that drives contact rates between susceptible and infected individuals.

Despite the fact that mobility data are often integrated into disease transmission models^3,6,7,10,18^, little is known about how changes in the spatial resolution of data can impact modeled results. Past research has explored various mobility spatial scales using data describing high-income county commuting patterns and mobility inferred from mobile phone data^16,19^, however similar studies in low or middle-income countries (LMICs) to date have rarely focused on this aspect of aggregation ^3,20^, with the exception of recent work in Ghana.^21^ Here, we explore the importance of the spatial scale of mobility to modeled disease transmission in a unique LMIC setting. Using mobility inferred from mobile phone data in Sri Lanka, we investigate the impact of increasing the granularity of mobility data on simulated epidemic dynamics. We find that that travel is heterogeneous amongst disaggregated data, with subunit urbanicity and proximity to unit borders determining deviance from average unit travel. These deviances impacted modeling results, where differences in epidemic magnitude and spatial invasion timing between models with varied mobility scales were highly reliant on the epidemic seeding location as well as disease transmissibility.

## Results

### Broader patterns of travel are consistent across spatial scales with weaker relationships for more granular data

We analyzed trip counts inferred from mobile phone data between three increasingly finer spatial scales (from coarsest: province, then district, to finest: division; **Figures 2a, d, g**). Missing travel estimates for origin-destination pairs were only observed at the division scale, accounting for 1.78% of the over 100,000 total routes. Missingness could either be a result of zero travel occurring between two locations or fewer than 5 daily trips occurring between two locations due to data censoring requirements. Overall, missingness corresponded with population density and trip distance where data were more likely to be missing if the origin or destination populations were small and the physical trip distance was large (**Table S.1**). In the observed data, the proportion of average daily trips remaining within the origin unit, or the internal trip proportion, generally decreased as the spatial scale became finer (**Figures S.1a, b, c, and Figures 2c, f, i**). For example, the average daily internal trip proportion at the province scale was 0.88 compared to 0.77 at the district scale and 0.23 at the division scale. Within each spatial scale, these internal trip proportions were generally highest in locations with larger populations (**Figures S.1d, S.1e**).

**Figure 2.**
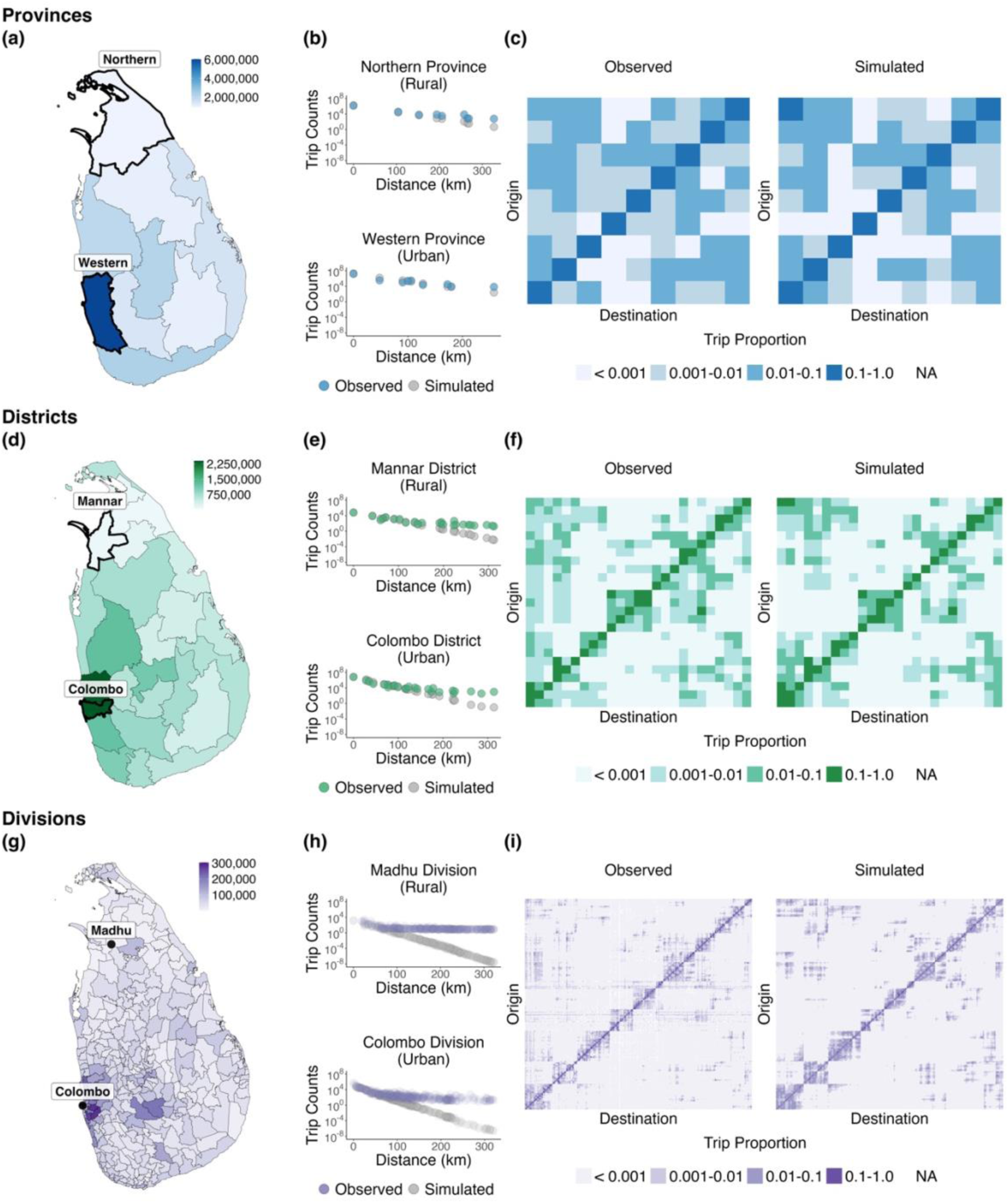
Population distributions, mobility trends, and trip proportion matrices remain consistent across spatial scales **(a), (d), (g)** Population distribution maps are displayed for provinces, districts, and divisions with the localities of Colombo and Madhu highlighted (including aggregated counterparts). **(b), (e), (h)** Mobility trends between trip distance and the number of trips for a rural (Madhu) and urban (Colombo) locality are shown at the province, district, and division scale and for observed and simulated data. **(c), (f), (i)** Trip proportion matrices for observed and simulated data at the province, district, and division scales are presented where darker colors correspond to higher trip proportions.

Regardless of the spatial scale, the total number of observed trips had a negative relationship with distance, although the strength of this relationship was the weakest for the finest spatial scales (**Table S.2**). This relationship held across origin locations including more urban, populated areas like Colombo as well as rural regions such as Madhu in the northern part of the country (**Figures 2b, e, h)**. Similarly, there were increased trips to and from more populated locations, although the relationship was also weaker for more granular spatial units (**Table S.2**). To model these relationships, we fit exponential gravity models to the observed mobility data and simulated expected trip counts at each level of spatial aggregation. While the agreement between simulated and observed trip counts was strong (correlation coefficients of 0.99 for province, 0.97 for district, and 0.81 for division levels), the accuracy and fit of these models was poorest for finer spatial scales (**Table S.3 and Figure S.2**).

### Mobility was not uniformly distributed as data moved from coarse to fine spatial scales

Noticeably, travel at larger spatial scales was not evenly distributed amongst nested subunits. If travel were evenly distributed across the larger spatial scale, we would expect subunits to have similar external trip proportions as the unit. However, that was not the case across the country. For example, the proportion of outgoing trips originating from the Northern province that left the Northern province (daily average external province trip proportion) was 0.05 while the proportion of outgoing trips originating from the Madhu division (nested within the Northern province) that left the Northern province was 0.32 (**Figure 3b**). This resulted in a large external province trip proportion difference of-0.27, reflecting that certain nested subunits may have a higher or even lower proportion of the population leaving the unit compared to the unit as a whole. Across all nested divisions within the Northern province, the daily average external province trip proportions ranged from 0.02 to 0.69 (**Figure 3c**).

**Figure 3.**
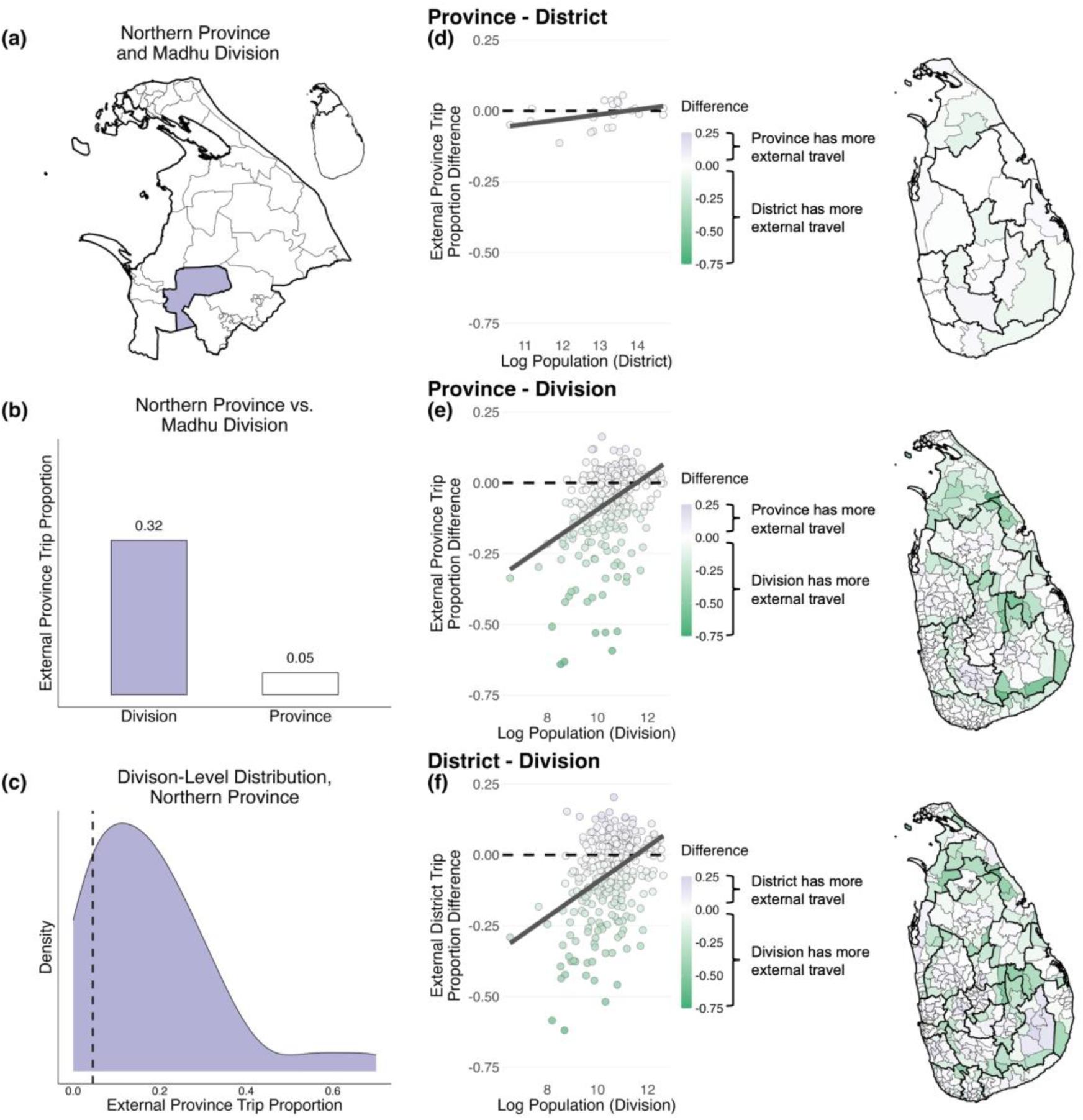
Mobility is not evenly distributed amongst the nested subunits of an encompassing unit **(a)** A map displaying divisions within the Northern province, with the Madhu division highlighted**. (b)** A comparison between the external province trip proportion of the Madhu division and the Northern province. **(c)** The distribution of external province trip proportions for all divisions within the Northern province. The dashed line displays the external province trip proportion for the Northern province. **(d)**, **(e)**, **(f)** Scatter plots with a linear trend line present the external trip proportion differences against the nested subunit population size for districts within provinces, divisions within provinces, and divisions within districts. Differences are calculated by subtracting the subunit proportion from the unit proportion. Maps display the external trip proportion differences spatially, where colors correspond to the scale that has more external travel.

On average, when comparing unit and subunit values (i.e. larger admin level locations with corresponding nested smaller admin level locations), the daily average external unit trip proportions differed by-0.06 (ranging from-0.64 to 0.16) for nested divisions within provinces. Average differences were similar for nested divisions within districts (-0.06, ranging from-0.62 to 0.20) and less extreme for nested districts within provinces (-0.01, ranging from-0.11 to 0.05) (**Figures 3d, e, f**). This external trip proportion difference tended to have larger negative values for subunits with smaller population sizes where more external unit travel occurred relative to the unit itself, although the strength of this relationship depended on the two spatial scales being compared (**Figure 3d**). As expected, subunits close to unit borders typically had higher values of external unit travel than interior subunits (resulting in a negative difference) likely due to the proximity and ease of travel to another unit (**Figures 3e, f**).

We also found that individuals commonly traveled to another subunit but remained within their home unit, clustering mobility that would not be captured in unit-aggregated data. For instance, divisions within the same province tended to have higher proportions of travel between each other than with divisions in other provinces. On average, the proportion of trips traveling to a division in the same province was 0.42 higher than the proportion of trips traveling to a division in a different province. This statistic was similarly higher at 0.23 for divisions within districts and-0.003 for districts within provinces, where there was almost an equal probability of traveling within versus leaving a unit (**Figure S.3**).

### Mobility data spatial scale impacted the modeled spatial invasion timing and epidemic magnitude of a theoretical disease outbreak

To explore how differences in mobility patterns for nested spatial scales impacted modeled disease dynamics, we ran multiple simulations of a theoretical outbreak with varying transmissibility parameters and initial infected locations (see **Materials and Methods**). For each spatial scale, coupling between locations was determined by the corresponding mobile phone data or simulated mobility from an exponential gravity model fit to the data (see **Supplementary Information**). We considered two scenarios with initial introduction events occurring in either the highly populated capital city (Colombo), or a contrasting rural area in northern Sri Lanka (Madhu) across a range of disease transmission parameters. The initial introduction location had a noticeable impact on the spatial disease dynamics between nested geographic scales when considering the division (admin 3) versus the district (admin 2) and province (admin 1) scales.

When comparing the two introduction location scenarios with fixed transmissibility parameters, we found that the spatial arrival timing for a theoretical disease outbreak depended on the spatial scale of the model in addition to the introduction location. For a disease introduced into Colombo, the median simulation estimated that infection was introduced into all 9 provinces in 66, 68, and 64 days for division, district, and province level models, respectively (**Figure 4c**). For a Madhu introduction, infection was introduced into all provinces in 59, 66, and 68 days for division, district, and province level models (**Figure 4f**). In general, larger scale models produced a faster epidemic when disease was initially introduced into Colombo. However, when a pathogen was seeded in the more remote area of Madhu, spatial dispersion varied slightly more between scales and the ordering of scale speed flipped. Here, the division scale model estimated faster arrival times than the coarser district and province models overall. Conversely, epidemic occurrence, magnitude, peak size, and peak timing all varied depending on the spatial scale of the model but not by the introduction location. For both Colombo and Madhu introduction event scenarios, the proportion of simulations that resulted in more than 100 total cases increased as the model spatial scale increased (**Figures 4a, d**). For instance, the Colombo introduction scenario had the epidemic proportion rise from 0.46 to 0.60 as the spatial scale increased from division to province. Epidemic magnitude and peak size generally increased while peak time decreased as the model spatial scale increased (**Figures 4b, e)**. For example, the final epidemic magnitude increased from 10.8 million to 12.5 million cases as the spatial scale increased from division to province for the Madhu introduction scenario. Simulated mobility data were about to create similar trends across spatial scales in epidemic occurrence, magnitude, peak size, and peak timing, however spatial invasion timing was not consistent with observed mobility data (**Figure S.5**)

**Figure 4.**
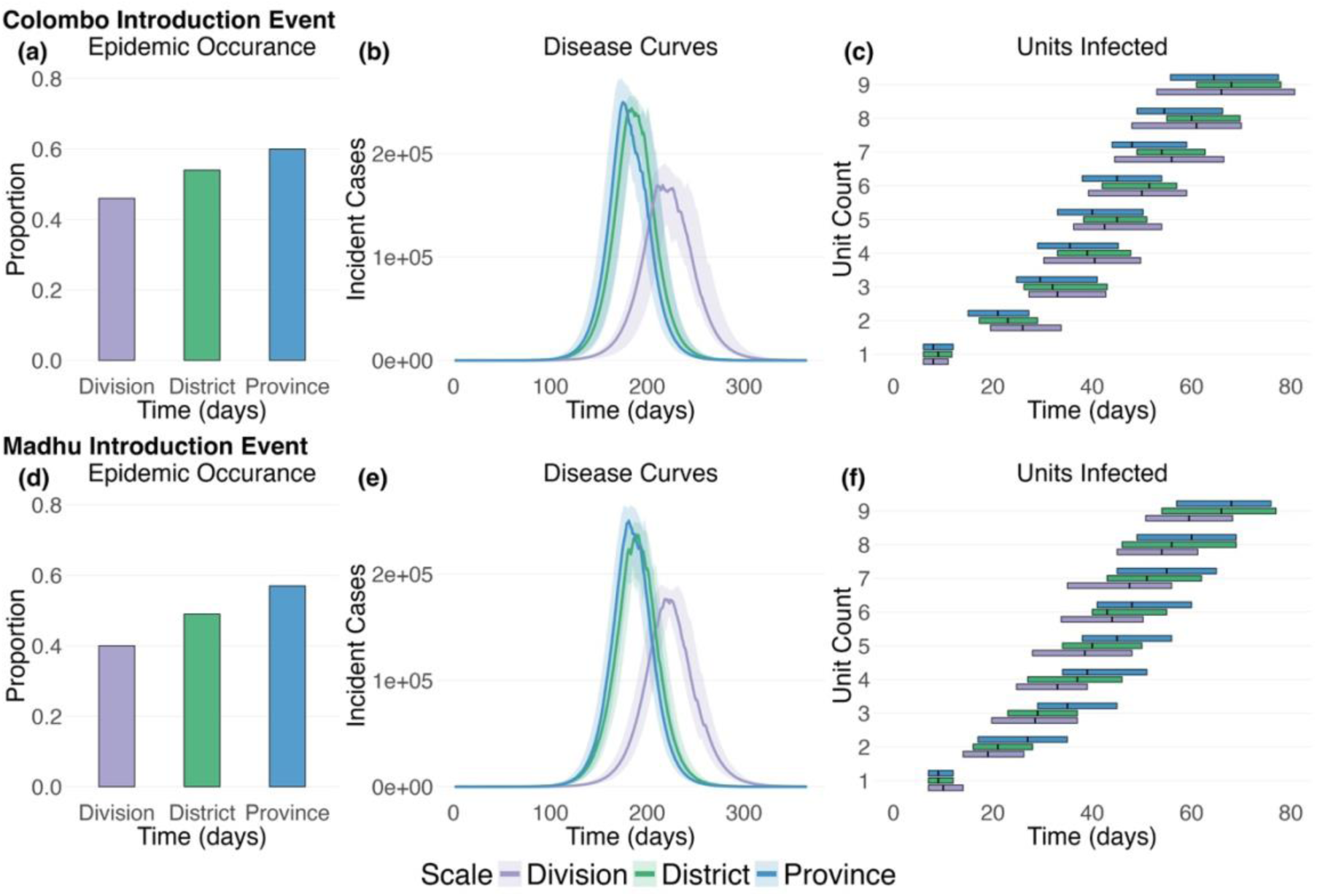
Initial epidemic seeding location and the spatial scales compared alter spatial arrival times, peak times, and peak magnitude Plots summarize two epidemic simulation scenarios: Disease dynamics modeled with observed mobility considering a Colombo introduction event and disease dynamics modeled with observed mobility considering a Madhu introduction event. **(a), (d)** Bar plots display the proportion of simulations that produced more than 100 total cases (i.e. resulted in an epidemic). **(b), (e)** Epidemic curves display the 50^th^, 25^th^, and 75^th^ percentiles of incident cases across the epidemic simulations for the entire country comparing the three different spatial scale models. **(c), (f)** Horizontal box plots show the timing and number of provinces infected as disease moves throughout the population examining the three different spatial scale models (aggregated to the province level for comparison). The 50^th^, 25^th^, and 75^th^ percentiles for when more than 1 cumulative case occurred in each province is displayed across simulations. 𝑅_0_ was set at 2, the latent period was set at 3 days, and the infectiousness duration was set at 5 days for each simulation.

We then explored model results where the three spatial scales were compared to one another in a pairwise fashion while changing one parameter: either the initial seeding location, latent period, transmissibility (R_0_), or duration of infectiousness (**Figure 5**). Boxplots describe the spread of the difference in results, with each dot representing the average difference between two models across 100 iterations. For scenarios that did not vary the initial seeding location, two different seeding locations (Colombo and Madhu) were compared to examine urban and rural dynamics. Across all potential epidemic seeding locations, units with smaller population sizes tended to produce faster but not necessarily larger epidemics with a fine-scale model compared to a larger-scale model (panel 1 of **Figures 5a, b**). This can be observed by the negative trends in **Figure 5a** and the relatively flat trends in **Figure 5b**. The consistent model differences in magnitude across seeding locations were likely due to the values of transmissibility parameters chosen.

**Figure 5.**
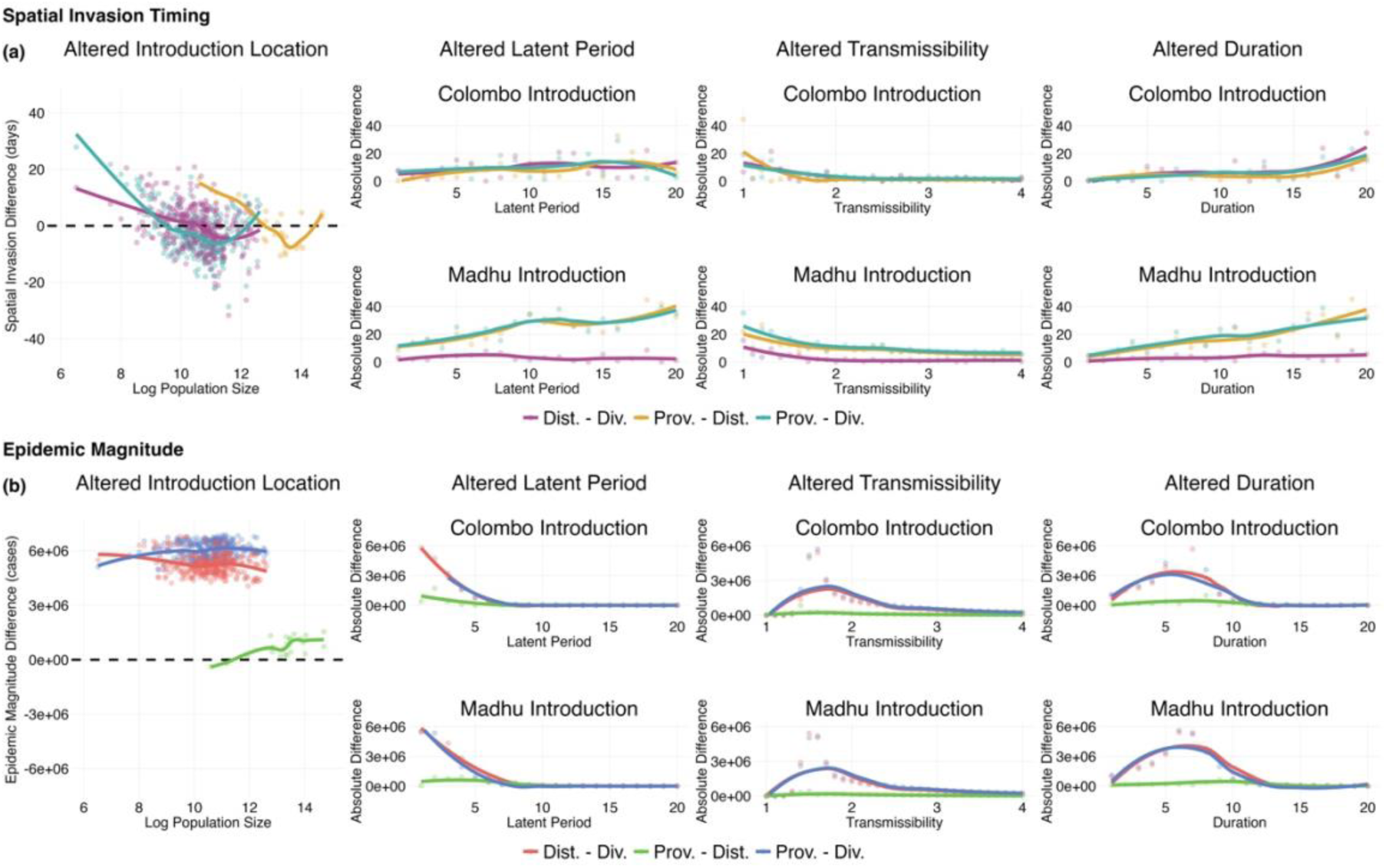
Differences in modeled spatial arrival times and epidemic magnitudes depend the on initial seeding locations, the scales compared, and pathogen transmissibility parameters **(a)** Points display the differences in average spatial arrival times in days between models run with various spatial scales of mobility data. Lines are the loess smoothed fit to the points. **(b)** Points display the differences in epidemic magnitude measured in cases between models run with various spatial scales of mobility data. Lines are the loess smoothed fit to the points. Each panel alters a single component of the simulation: initial seeding location, disease latent period, disease transmissibility, and disease duration of infectiousness. For more information see **Materials and Methods** and **Supplementary Information**.

In addition to the initial seeding location, the discrepancy between simulations at different scales also depended on certain transmissibility characteristics (panels 2, 3, and 4 in **Figures 5a, b**).

As disease transmissibility (measured as R_0_) increased, models at different spatial scales tended to produce similar estimates of spatial invasion timing and epidemic magnitude, indicated by the absolute difference trend lines approaching 0 as R_0_ increases to 4. When examining the disease latent period and duration of infectiousness, larger values of these parameters resulted in models with smaller differences in epidemic magnitude and generally larger differences in spatial invasion timing. However, these differences in spatial invasion timing were also guided by the epidemic seeding location. Simulations with simulated mobility data did not reproduce the same spatial invasion timing difference trends as models informed by the observed mobility data, where aggregated models with simulated data almost always resulted in a faster epidemic compared to finer scale models. For epidemic magnitude, large values of transmissibility parameters resulted in smaller differences between models run with simulated mobility data, similar to observed data (**Figure S.6**).

### Mobility and homogeneous mixing differences between scales drive modeling differences

Altering the spatial scale of an infectious disease model both changes the scale of mobility data that is incorporated into the model, but also the spatial scale of individuals that are assumed to be mixing homogeneously. Since both of these factors are simultaneously altered, it is difficult to attribute model differences to one component or the other. As a sensitivity analysis, mobility data were rescaled so the mobility patterns at smaller scales was adjusted to match mobility at larger scales. With the rescaled mobility data, the proportion of trips leaving the Northern province from Madhu division were scaled down from 0.32 under the original observed mobility data to 0.05, eliminating the differences in trip proportions between nested subunits and units (**Figure 3b, Figure S.7**). Building off **Figure 4**, **Figure S.8** examines similar scenarios but with rescaled mobility data. In general, while we do not observe many changes between observed and rescaled data, the epidemic occurrence proportion dropped for province level models and spatial invasion timing was noticeably slower for the district level model when considering epidemics seeded in Madhu. For rescaled data, aggregated models were more likely to create faster epidemics when compared to observed data (**Figure S.9a**). When examining epidemic magnitude, there were little differences between observed and rescaled data apart from province and division level model comparisons where rescaled data produced slightly smaller discrepancies in the total number of cases (**Figures S.9b).** Even when removing the impact of mobility data differences between scales, model differences persisted from altering the assumption of the scale of homogeneous mixing.

## Discussion

This work explores the impacts of different spatial aggregations of mobility data on a model of infectious disease transmission. Using detailed, spatially explicit mobility data inferred from mobile phone call data records in Sri Lanka, we observed differences in the types, frequency, and proportion of travel across increasingly coarser spatial scales. We then developed a spatial model of infectious disease transmission and explored how differences in the spatial scale of mobility data impacted the epidemic trajectory. Similarities between simulations using finer or coarser mobility data resolutions were dependent on the disease transmissibility parameters, the population size of the initial seeding location, the epidemic statistic itself, and the spatial scales being compared. Overall, we found that discrepancies in the estimated spatial invasion time of a disease for models at different spatial scales depended on the introduced location and transmissibility of the pathogen, similar to results from analyses in Ghana.^21^ Fine-scale models were likely to produce faster diffusion compared to large-scale models if the pathogen was introduced into a rural location while the opposite was true for urban seeding locations, although these differences were less pronounced for highly transmissible pathogens. While discrepancies in the modeled spatial invasion time between scales was dictated primarily by the seeding location, differences in epidemic magnitude were largely determined by transmissibility parameter values. Diseases with long latent periods, high transmissibility, or long durations of infectiousness all produced similar epidemic sizes across models with varying spatial resolutions. Although models run with simulated data did not replicate spatial invasion results from observed data, trends were consistent when comparing epidemic magnitude (**Figures S.5, S.6**).

Critical to this analysis is the consideration of not just changes to the spatial scale of mobility data incorporated into infectious disease models, but also the assumed scale of homogeneous mixing. Primary analyses examined each of these factors in tandem when altering the spatial scale of infectious disease simulation. However, in sensitivity analyses we aimed to isolate the impact of homogeneous mixing by rescaling mobility data to eliminate differences in travel between spatial resolutions. We observed that although the spatial scale of mobility data contributed to differences between models, discrepancies between models persisted when only changes to the spatial scale of homogeneous mixing was examined. As a result, future work should explore the appropriate spatial scale of mobility data that recreates observed disease dynamics in addition to the appropriate spatial scale of contact where homogeneous mixing occurs.

These results were limited in terms of data representativeness and model specifications. While mobile phone data had high geographic coverage, groups such as elderly adults, children, and those of lower socioeconomic status may not be included, which could reduce generalizability of the movement information.^22,23,24^ Despite the fact that travel from mobile phone data was available on fairly granular spatial scales (admin 3), these still only represent the approximate location of an individual subscriber. In addition, these data were censored such that data were only available if at least 5 trips were recorded in a day. This represents an underestimation of the true number of recorded trips between locations. Moreover, some simplifying assumptions were made in the transmission model. For example, we assumed homogenous mixing within each location in the metapopulation which ignores within location heterogeneities in movement and contact patterns.^25^ We also did not vary the proportion of the population who were susceptible, since we were using the model to illustrate differences by the granularity of the mobility data, which may not represent real world conditions. Overall, we did not consider additional heterogenies in susceptibility across the population or differential contact rates, beyond mobility patterns and homogeneous mixing. Thus, additional work should be conducted to explore how these factors may intersect with mobility patterns across spatial scales. Further, we only explored these patterns in a single setting using a single data source. While the generalizability of this work to other locations and data types may be impacted, we nonetheless provide a framework to explore these factors in other populations.

Recent work has shown that fine scales of mobility data reproduced the spatial diffusion of COVID-19 in the United States better than aggregated scales.^16^ However, additional research is needed to fully evaluate what scale of mobility data should be used for various questions. While the spatial scale of infectious disease models is often dictated by data availability, additional evaluations of the significance of scale are needed. In particular, when infectious disease models are being used to inform public health decisions, considering a wide range of diverse mobility data scales may be needed to ensure the robustness of final conclusions. Ideally, this would be informed by a mechanistic understanding of on what scales human behavior facilities disease transmission. As additional data sets become available that are able to measure increasingly granular movement and contact patterns, these types of studies may be feasible.

## Materials and Methods

### Geographic Data

Geographic shapefile data for provinces (administrative level 1), districts (administrative level 2), and divisional secretariats (administrative level 3) in Sri Lanka were obtained from the Humanitarian Data Exchange.^26^ Shapefiles contained polygon coordinates for 9 provinces, 25 districts, and 339 divisional secretariats. The number of divisional secretariats was inconsistent with the 331 designated by the Sri Lankan government^27^ due to the disaggregation of single units into two or three in the shapefile data. Additional information on administrative unit disagreement between datasets is provided in the supplement (**Tables S.5a and S.5b**).

Divisional secretariat shapefile data were aligned to the Sri Lankan government distinctions and later to divisional secretariats in the mobile phone data.

### Population Data

Data on population counts were downloaded from WorldPop.^28^ Population data, the 2020 constrained United Nations (UN) adjusted data at the 100m resolution level, were used to estimate population totals for administrative units at each spatial scale. Every tile was adjusted by the ratio of Sri Lanka’s 2021 UN population to Sri Lanka’s 2020 UN population^29^ so totals reflected 2021 population estimates, the most recent data at the time of analysis. After adjustment, all tiles were assigned to a province, district, and divisional secretariat by performing a spatial join in ArcGIS.^30^ Tile centroids that fell within the bounds or were closest to an administrative unit were assigned to the corresponding unit. Aggregated population data are displayed in **Figure 2**.

### Mobile Phone Data

Mobile phone data was provided in the form of aggregated and anonymized call data records (CDRs) from a leading telecommunications operator in Sri Lanka, Dialog Axiata. Based on financial documentation, the company had over 17.7 million mobile subscribers across all coverage networks in 2021.^31^ Call data records are able to document a subscriber’s movement when calls or short messaging service (SMS) texts made by the subscriber are routed through the nearest cellular tower. Approximate locations of subscribers are then estimated by obtaining location information of each cellular tower in the network.^14^ Subscriber movement in this dataset was aggregated to the divisional secretariat (administrative level 3) by the data distributor.

Mobile phone data captured the number of daily trips between two unique locations, meaning that it was possible for highly mobile subscribers to be included in multiple origin-destination pairs each day. Data were available from November 1, 2021 until March 15, 2022.

Mobile phone data were censored if five or fewer subscribers traveled on a route each day. Data missingness was evaluated using univariate logistic regression, with missingness as a binary outcome and origin population, destination population, and trip distance as explanatory, continuous variables. In subsequent analyses, missing trip count information in mobile phone data due to censoring was set to zero.

### Calculating Trip Proportions

Mobile phone data were aggregated and averaged across time to create daily average trip counts for each origin-destination pair at the division, district, and province scales. Trip proportion (𝑝) was calculated as the fraction of trips (𝜏) to a destination (𝑗) out of all trips from a common origin (𝑖):

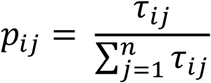

Origin-destination matrices were created from these trip proportions for each administrative level, resulting in a total of three origin-destination matrices. Trip proportions along with the proportion of trips staying within an administrative unit were compared across spatial scales. Additionally, external trip proportions were compared between units and disaggregated subunits at all combinations of spatial scales.

### Fitting Mobility Models to Mobile Phone Data

Exponential gravity models were fit to each spatial scale of mobile phone data using the ‘mobility’ package in R, which uses Bayesian inference and Monte Carlo Markov Chain (MCMC) methods to fit parameters.^32^ Estimated trip counts (𝜏_𝑖𝑗_) were simulated based on the origin population (𝑁_𝑖_), destination population (𝑁_𝑗_), and distance in kilometers between origin and destination (𝑑_𝑖𝑗_).

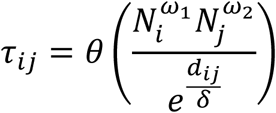

Where 𝜃 is a proportionality constant, 𝜔_1_ and 𝜔_2_ weight the importance of the origin or destination population, and 𝛿 serves as a non-negative deterrence factor on the trip distance. Each parameter has an uninformative gamma prior, with the exception of 𝛿 which employs a normal distribution prior truncated at 0.^33^

### Simulating Infectious Disease Dynamics

Disease dynamics at different spatial scales were simulated using discrete-time stochastic metapopulation SEIR models with corresponding levels of spatial granularity (see **Supplementary Information**).^34,35^ The force of infection was calculated by using mobility data to account for individuals staying in their patch, visitors traveling to other patches, and residents returning to their home patch based on.^16^ We simulated daily disease dynamics for a year with a single infected individual in an initial location.

Ranges of transmission parameters were explored to examine potential modifications of model result differences across mobility data spatial resolutions. For each modelling scenario at a given spatial resolution and combination of seeding location, 𝑅_0_ (1-4), latent period (1-20 days), and infectiousness duration (1-20 days), 100 simulations were run to estimate the average number of incident cases for each unit at all time steps. Epidemic magnitude and spatial invasion statistics were then calculated from the average result across simulations.

### Sensitivity Analysis: Rescaling Trip Proportions to Evaluate Homogeneous Mixing

Previous models compared changes between the scale of mobility data in addition to the geographic scale of the model, which in turn altered the number of individuals assumed to be homogeneously mixing. To examine modeling sensitivity to spatial scale alterations independent of differences in observed mobility, finer-scale trip proportion matrices were rescaled to match mobility patterns at the larger-scale matrix. For instance, when comparisons were between district and province scales, district mobility was rescaled such that province travel at the district scale was the same as province travel at the province scale. By rescaling, the only differences between district and province trip proportion matrices were the number of metapopulation patches and not the connectivity between patches. Modeling and result comparisons were performed as stated above and in the supplement. This analysis was performed to assess how much the spatial scale of homogeneous mixing contributed to model result differences.

## Code and Data Availability

All analyses were performed in R version 4.2.1^36^ and ArcGIS Pro version 2.9.^30^ The code used to analyze mobility data, run infectious disease models, and produce all figures is located on GitHub.^37^ Geographic boundary and population data are all publicly available through the indicated sources. While original mobility data are not permitted to be shared, simulated mobile phone data from gravity models are provided on GitHub.

## Data Availability

Data is proprietary but simulated data is made publicly available.

https://github.com/rcorgel/mobility-spatial-scale/tree/main/simulated%20data

## Acknowledgments

We would like to thank Dulakshi Abeygunawardhana, Fiona Alexander, Maheeka Kumarihamy, and Pranithye Gunasekara for data collection and entry relevant to this project.

