## Supplementary Material for "Characterizing the impact of incorporating spatially aggregated human mobility data into infectious disease models"

### Metapopulation Model Mechanics

In the mechanistic model used to simulate disease dynamics, individuals transitioned between susceptible, exposed, infected, and recovered compartments, analogous to the chronological transition of humans between disease states. The metapopulation model accounted for spatial disease dynamics by treating each geographic unit as its own patch. The number of patches fluctuated depending on the spatial resolution scale of the model. For instance, the district-scale metapopulation model contained 25 patches, equivalent to the 25 districts in Sri Lanka. The equations of each compartment in the model are listed below for patch $i$ at time $t$ are:

$$S_{i,t} = S_{i,t-1} - \epsilon_{i,t}$$
$$E_{i,t} = E_{i,t-1} + \epsilon_{i,t} - \iota_{i,t}$$
$$I_{i,t} = I_{i,t-1} + \iota_{i,t} - \rho_{i,t}$$
$$R_{i,t} = R_{i,t-1} + \rho_{i,t}$$

Where $S$ is the number of susceptible individuals, $E$ is the number of exposed individuals, $I$ is the number of infected individuals, $R$ is the number of recovered individuals, $\epsilon$ is the number of new exposures, $\iota$ is the number of new infections, and $\rho$ is the number of new recoveries. Frequency dependent disease transmission, untethered from population size, was assumed to isolate the effect of mobility data spatial aggregation on modeling results. Travel was incorporated into the model as an origin-destination matrix containing trip proportions for the specified spatial resolution.

### Mobility-Informed Force of Infection Calculation

Mobility data was used to calculate the force of infection for a given patch and time step. Infected individuals were assumed to have similar travel patterns as both susceptible and recovered individuals. Thus, the force of infection accounted for infected individuals staying in their patch ($s$), visiting other patches ($v$), and returning to their home patch ($r$).[1]

The equation for the force of infection ($\lambda$) in patch $i$ at time $t$ is as follows:

$$\lambda_{i,t} = \lambda_{ii,t}^{s} + \sum_{i \neq j} \lambda_{ji,t}^{v} + \sum_{i \neq j} \lambda_{ij,t}^{r} \qquad with \begin{cases} \lambda_{ii,t}^{s} = \beta p_{ii,t}^{2} \frac{I_{i,t}}{\widehat{N_{i,t}}} \\ \lambda_{ji,t}^{v} = \beta p_{ii,t} p_{ji,t} \frac{I_{j,t}}{\widehat{N_{i,t}}} \\ \lambda_{ij,t}^{r} = \beta p_{ij,t} \frac{\widehat{I_{j,t}}}{\widehat{N_{j,t}}} \end{cases}$$

Where $N$ is the population size of a patch (the sum of $S$, $E$, $I$, and $R$ compartments), $\beta$ is the transmissibility parameter, and $p$ is the trip proportion of either staying in a location ($ii$), visiting a destination ($ij$), or returning from a destination ($ji$). The effective population and the effective number of infections are defined as:

$$\widehat{N_{i,t}} = p_{ii}N_i + \sum_{i \neq j} p_{ji}N_j$$

$$\widehat{I_{i,t}} = p_{ii}I_i + \sum_{i \neq j} p_{ji}I_j$$

**Simulating the Stochastic Nature of Travel, Exposure, Infection, and Recovery**

Prior to calculating the force of infection for each time step, the number of trips between and within locations were drawn from a binomial distribution based on the population size and average daily travel patterns then normalized to travel proportions. This allowed for a random travel element to be included in the model:

$$\tau_{ij} \sim Binomial(N_i,\ p_{ij})$$

Where $\tau_{ij}$ is the number of trips from an origin ($i$) to a destination ($j$), $N_i$ is the population size of the origin location, and $p_{ij}$ is the average daily travel proportion for the origin-destination pair calculated from observed data. This was done for all origin-destination pair combinations.

Similar to travel, exposure, infection, and recovery were also allowed to vary in a stochastic nature. New exposures ($\epsilon$) were drawn from a binomial distribution based on the number of susceptible individuals in the previous time step ($S$) and the probability of exposure which depends on the force of infection ($\lambda$). New infections ($\iota$) were drawn from a binomial distribution based on the number of exposed individuals in the previous time step ($E$) and the probability of infection which depends on the latent period of disease ($\frac{1}{\sigma}$). Finally, new recoveries ($\rho$) were also drawn from a binomial distribution based on the number of infected individuals in the previous time step ($I$) and the probability of recovery which depends on the disease duration ($\frac{1}{\gamma}$).

$$\epsilon_{i,t} \sim Binomial(S_{i,t-1}, 1 - e^{-\lambda_{i,t}})$$
$$\iota_{i,t} \sim Binomial(E_{i,t-1}, 1 - e^{-\sigma})$$
$$\rho_{i,t} \sim Binomial(I_{i,t-1}, 1 - e^{-\gamma})$$

**Metapopulation Simulation Specifics**

Disease dynamics produced by metapopulation models were examined on a daily time step, for 365 days, following the introduction of a single individual infected with a novel pathogen into Sri Lanka. Based on the model structure and brief time-period explored, recovered individuals were assumed to be immune, ineligible for reinfection. In these model simulations, the proportion of the population susceptible was set at 0.90 for all metapopulation patches, reflecting a population with little existing immunity to infection. Births and deaths were not factored into the model, assuming a constant population size over the year-long period. Additionally, the model assumed homogeneous mixing within each patch.

To examine changes in infectious disease modelling results between different mobility data spatial resolutions, simulations of the metapopulation model were carried out at the province, district, and divisional secretariat scales. A wide range of epidemic scenarios were explored across simulations. All potential seeding locations, $R_0$ values from 1 to 4, latent periods from 1 to 20 days, and durations of infectiousness from 1 to 20 days were investigated. Parameter ranges were chosen as reasonable values for an acute respiratory infection of pandemic potential.[2,3,4,5] As disease was simulated across a range of values for one parameter, the remaining parameters were held constant.

Once epidemic scenarios were simulated, epidemic magnitude and spatial invasion timing were measured to compare between model runs. Epidemic magnitude was calculated as the sum of incident infections that occurred throughout the year-long time-period. Spatial invasion timing, on the other hand, was estimated as the average time across simulations at which all units had at least one cumulative case (excluding the introduction unit). Epidemic magnitude and spatial invasion timing were the primary statistics compared between model scenarios.

**Supplementary Figures and Tables**

**Table S.1. Missing data univariate regression results by explanatory variable for observed division scale (administrative level 3) mobility data**

| Univariate Explanatory Variable | Spatial Scale | Coefficient | P-Value |
|---|---|---|---|
| Origin Population | Division | -3.804 | <2.00E-16*** |
| Destination Population | Division | -4.907 | <2.00E-16*** |
| Trip Distance | Division | 0.024 | <2.00E-16*** |

**Significance.** 0 '***' 0.001 '**' 0.01 '*' 0.05 '.' 0.1 ' ' 1
**Notes.** Population variables are scaled such that coefficients represent change in log odds of missingness per change in 100,000 people. The distance variable is scaled so coefficients represent change in log odds per change in 100 km trip distance. Administrative levels 1 and 2 are not present for mobile phone data since no missingness was observed in those datasets.

**Figure S.1. Relationships between the internal trip proportion of a unit and unit population by mobility data type and spatial scale**

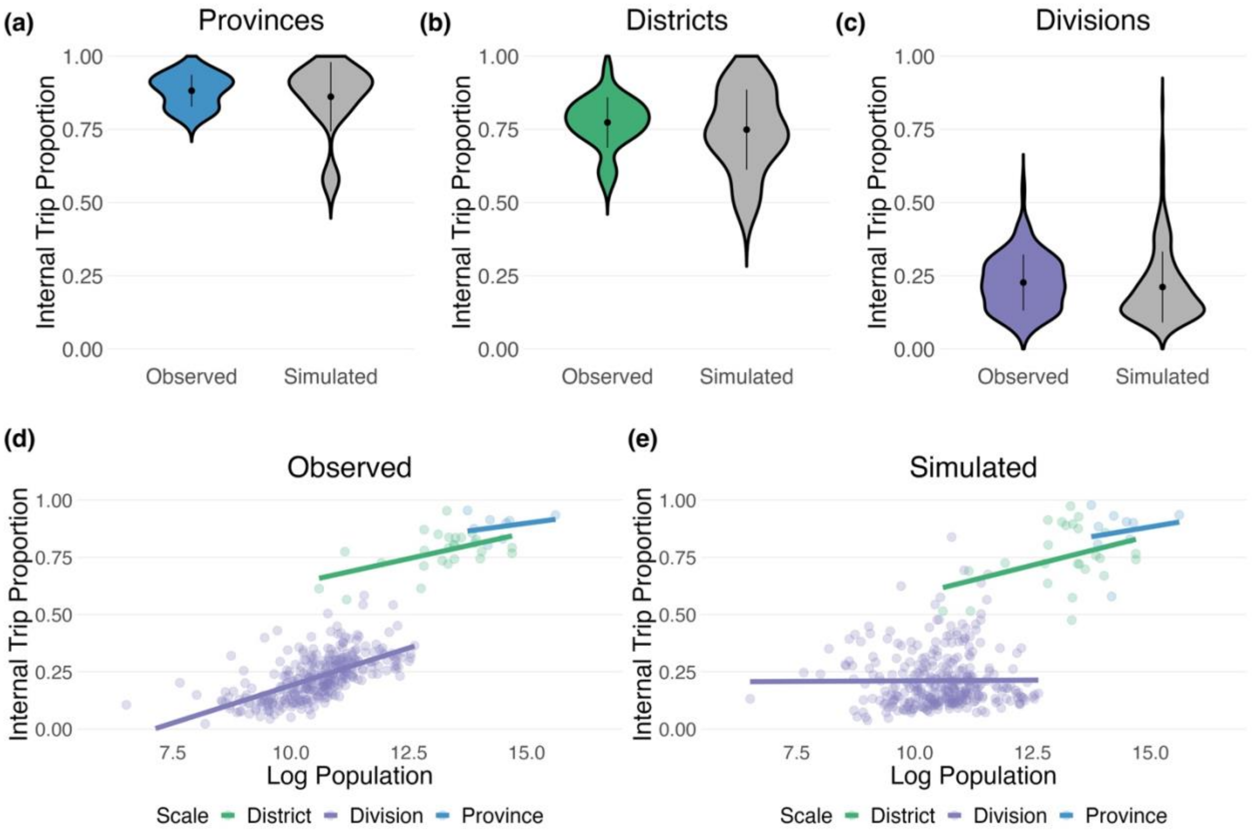


**Notes. (a), (b), (c)** Daily average internal trip proportion distributions for units at the province, district, and division scale stratified by observed and simulated mobility data. **(d), (e)** The relationship between a unit's daily average internal trip proportion and its log population size for divisions, districts, and provinces for observed and simulated mobility data.

**Table S.2. Correlation of trip count between distance, origin population, and destination population by mobility data type and spatial scale**

| Spatial Scale | Data Type | Distance (km) | Origin Population | Destination Population |
|---|---|---|---|---|
| Province | Observed | -0.42 | 0.25 | 0.25 |
| | Simulated | -0.42 | 0.26 | 0.27 |
| District | Observed | -0.27 | 0.18 | 0.18 |
| | Simulated | -0.29 | 0.19 | 0.20 |
| Division | Observed | -0.13 | 0.10 | 0.10 |
| | Simulated | -0.22 | 0.17 | 0.17 |

**Notes.** All results display Pearson correlation coefficients between the number of trips per route and the variable of interest.

**Table S.3. Gravity model parameter fits and DIC by spatial scale**

| Spatial Scale | Delta ($\delta$) | Omega 1 ($\omega_1$) | Omega 2 ($\omega_2$) | Theta ($\theta$) | DIC |
|---|---|---|---|---|---|
| Province | 24.80 | 0.48 | 0.84 | 0.02 | 4.39E+06 |
| District | 18.05 | 0.48 | 0.82 | 0.03 | 8.04E+06 |
| Division | 12.38 | 0.56 | 0.57 | 0.08 | 4.76E+07 |

**Notes.** Exponential gravity models were fit to each spatial scale of mobile phone data. More information on the model and process can be found in the **Materials and Methods** section.

**Figure S.2. Trip count and trip proportion comparisons between observed and simulated mobility data**

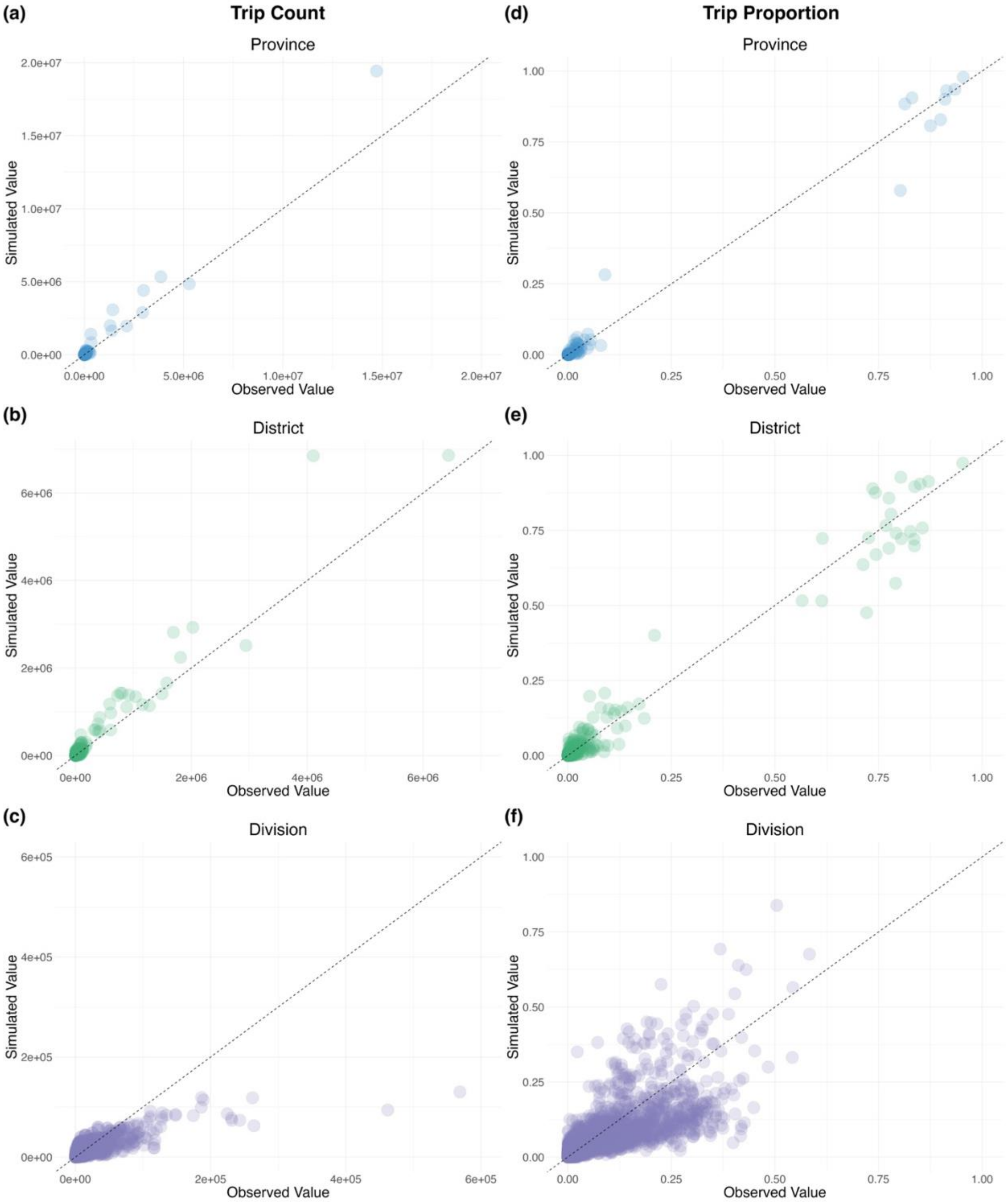


**Notes. (a), (b), (c)** Correlations of 0.99, 0.97, and 0.81 between observed and simulated trip counts at the province, district, and division levels, respectively. **(d), (e), (f)** Correlations of 0.99, 0.98, and 0.82 between observed and simulated trip proportions at the province, district, and division levels, respectively.

### S.3 Trip Proportion Comparison between Nested Units with Simulated Mobility Data

#### Province - District

(a)

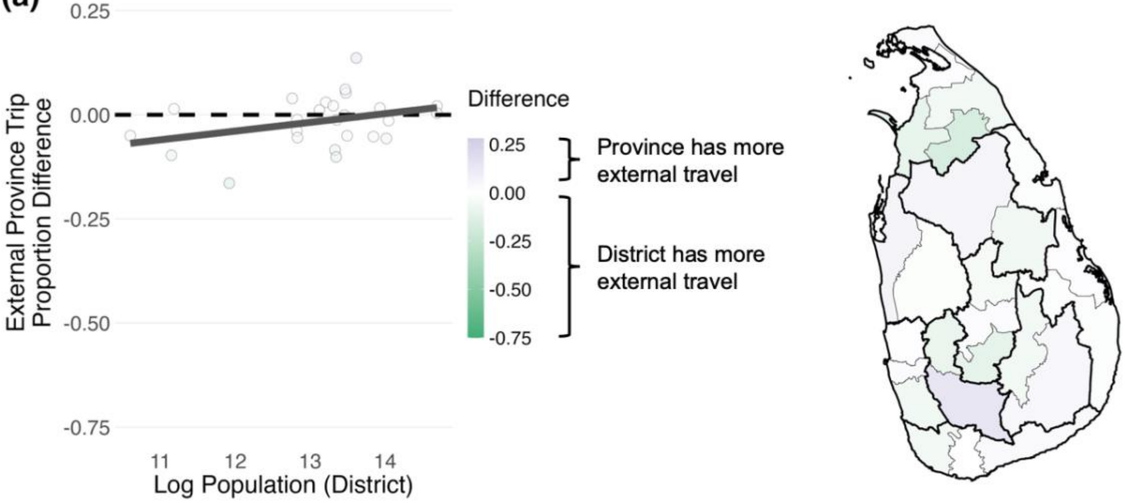


#### Province - Division

(b)

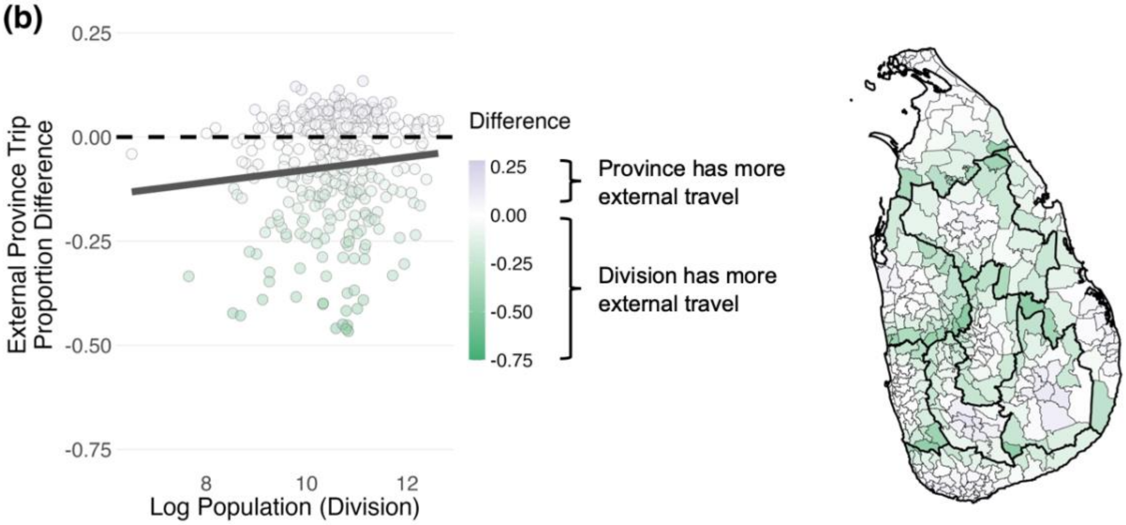


#### District - Division

(c)

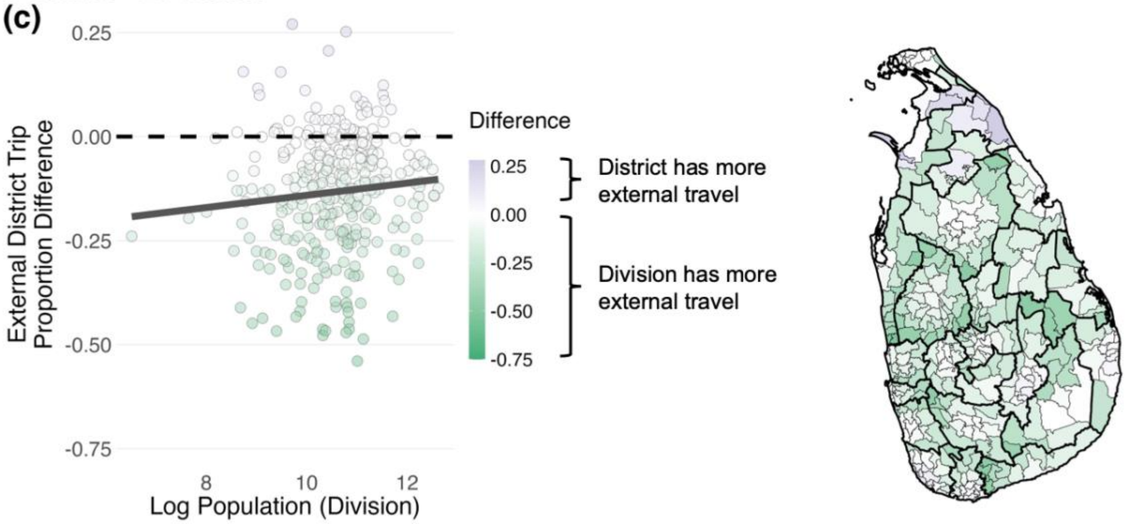

**Notes. (a)**, **(b)**, **(c)** Scatter plots with a linear trend line present the external trip proportion differences against the nested subunit population size for districts within provinces, divisions within provinces, and divisions within districts. Differences are calculated by subtracting the subunit proportion from the unit proportion. Maps display the external trip proportion differences spatially, where colors correspond to the scale that has more external travel.

**Figure S.4. Mobility clustering between nested subunits within a unit by nested spatial scales**

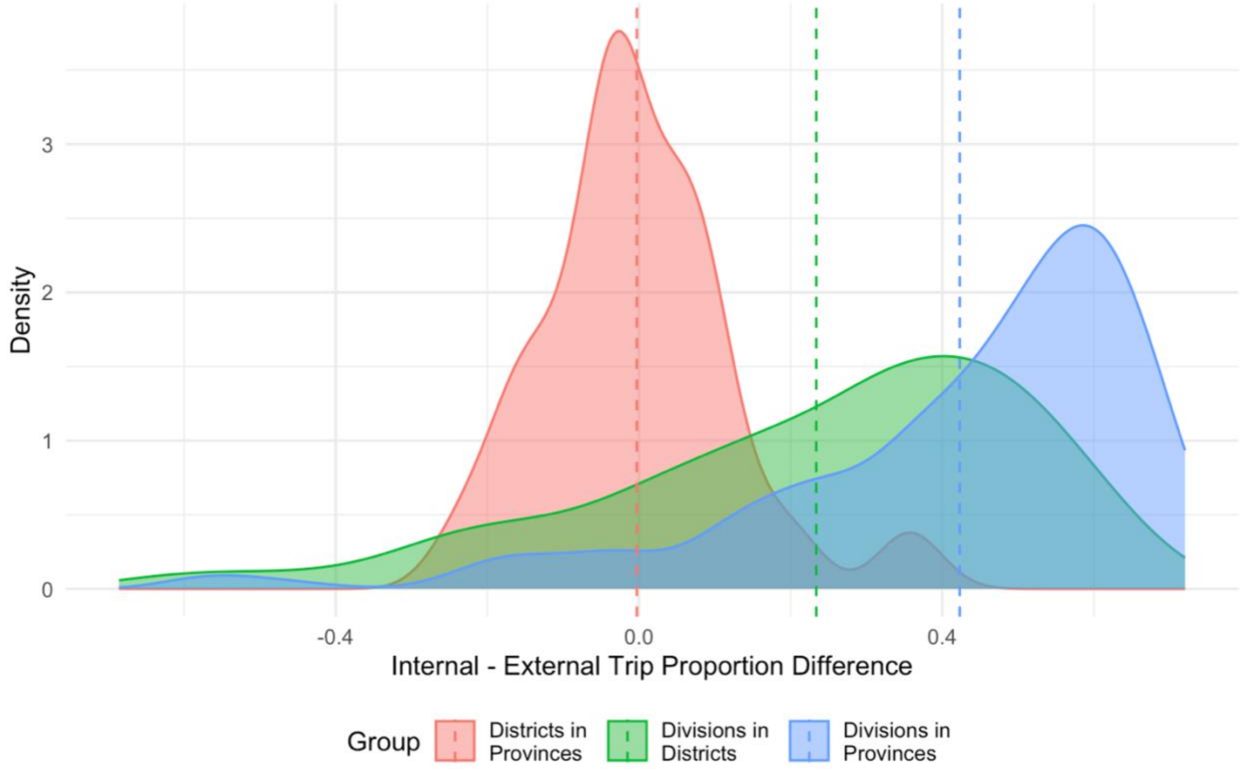


**Notes.** The distribution of the difference between a nested unit's proportion of trips to another unit within the same encompassing unit (internal) verses to a unit outside of the encompassing unit (external) is shown for various nested spatial scales. Positive values indicate clustering of travel where there is a higher proportion of travel to another subunit within an encompassing unit rather than an external unit.

### S.5 Epidemic Occurrence, Disease Curves, and Spatial Diffusion of Modeled Epidemics with Simulated Mobility Data

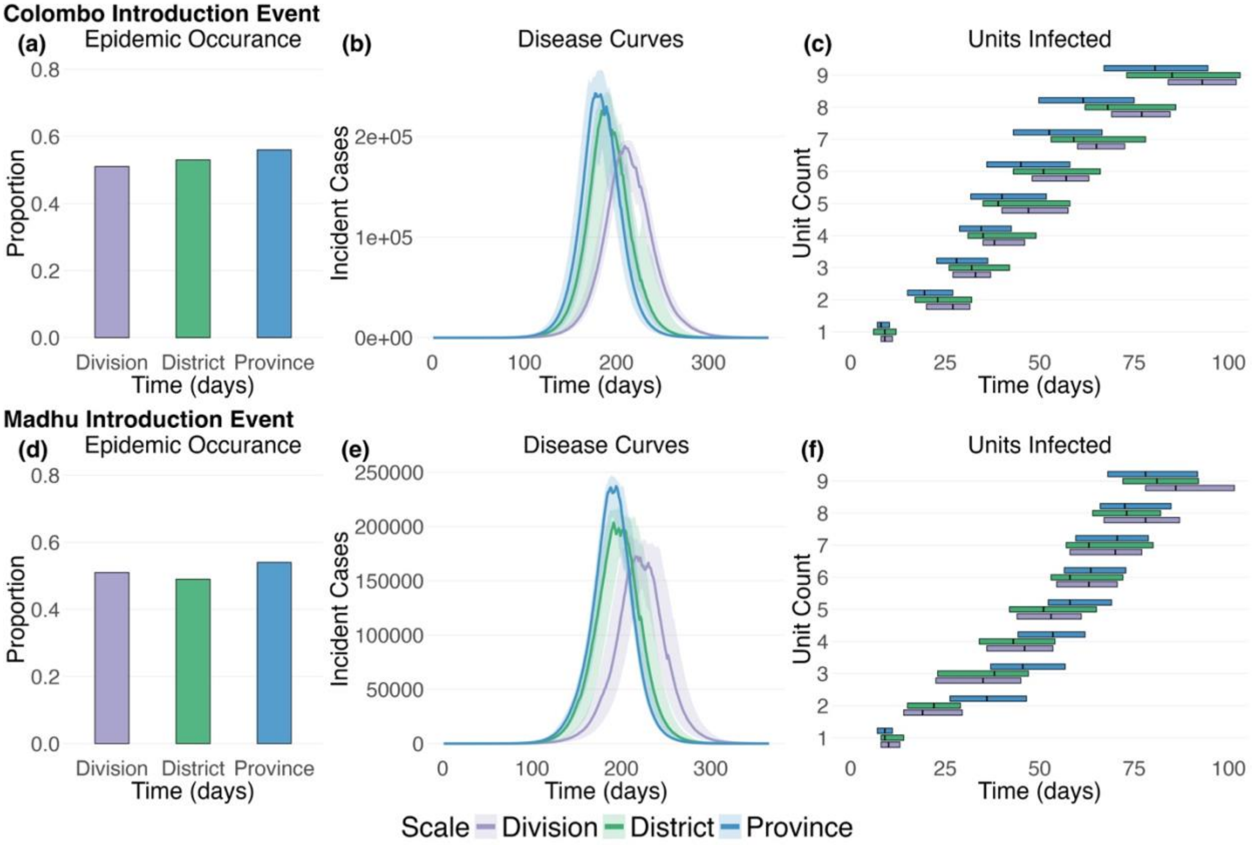


**Notes.** Plots summarize two epidemic simulation scenarios: Disease dynamics modeled with simulated mobility considering a Colombo introduction event and disease dynamics modeled with simulated mobility considering a Madhu introduction event. **(a), (d)** Bar plots display the proportion of disease simulations that produced more than 100 total cases (i.e. resulted in an epidemic). **(b), (e)** Epidemic curves display the 50$^{th}$, 25$^{th}$, and 75$^{th}$ percentiles of incident cases across the epidemic simulations for the entire country comparing the three different spatial scale models. **(c), (f)** Horizontal box plots show the timing and number of provinces infected as disease moves throughout the population examining the three different spatial scale models (aggregated to the province level for comparison). The 50$^{th}$, 25$^{th}$, and 75$^{th}$ percentiles for when more than 1 cumulative case occurred in each province is displayed across simulations. $R_0$ was set at 2, the latent period was set at 3 days, and the infectiousness duration was set at 5 days for each simulation.

### S.6 Pairwise Differences in Epidemic Statistics between Models Run at Various Spatial Scales with Simulated Mobility Data

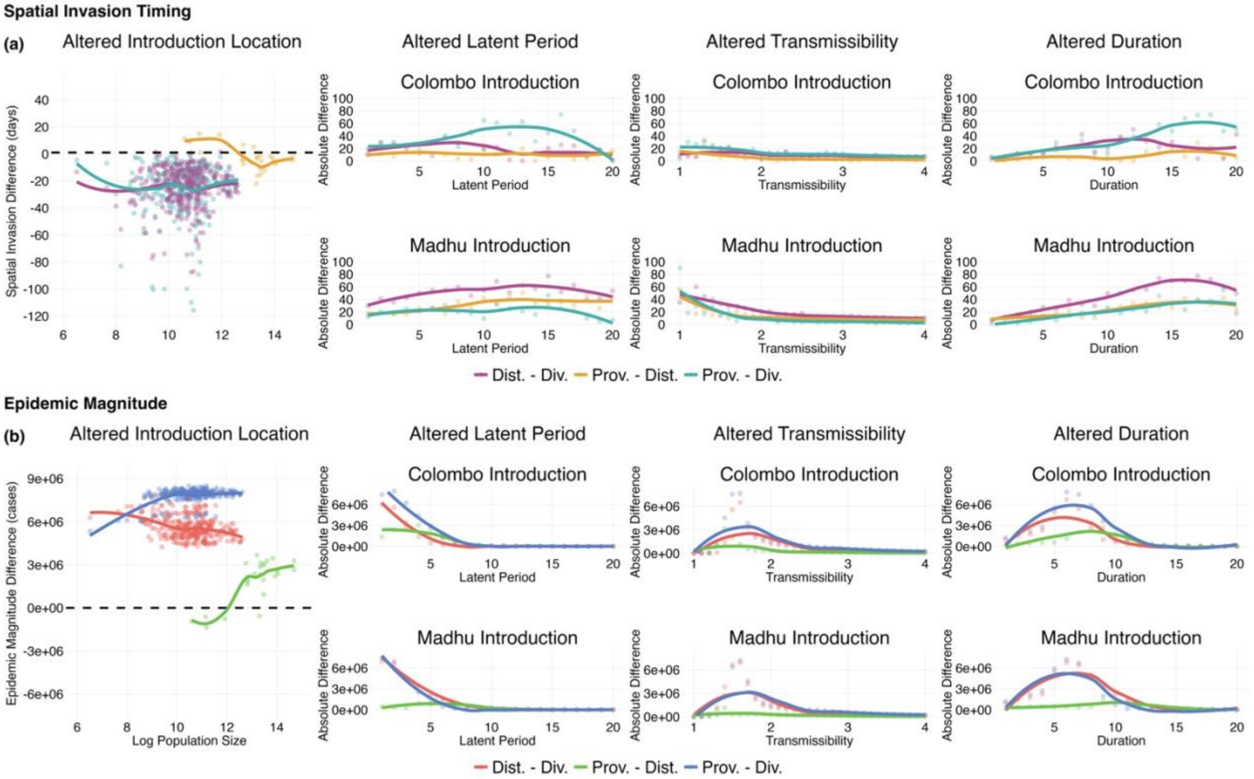


**Notes. (a)** Points display the differences in average spatial arrival times in days between models run with various spatial scales of mobility data. Lines are the loess smoothed fit to the points. **(b)** Points display the differences in epidemic magnitude measured in cases between models run with various spatial scales of mobility data. Lines are the loess smoothed fit to the points. Each panel alters a single component of the simulation: initial seeding location, disease latent period, disease transmissibility, and disease duration of infectiousness.

**S.7 Trip Proportion Comparison between Nested Units with Rescaled Mobility Data**

### Province - District

**(a)**

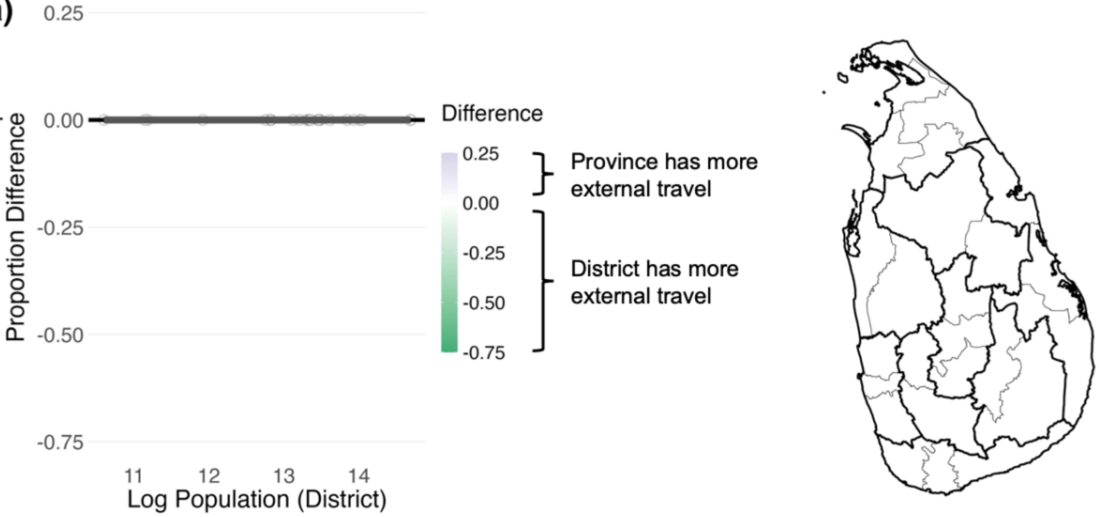


### Province - Division

**(b)**

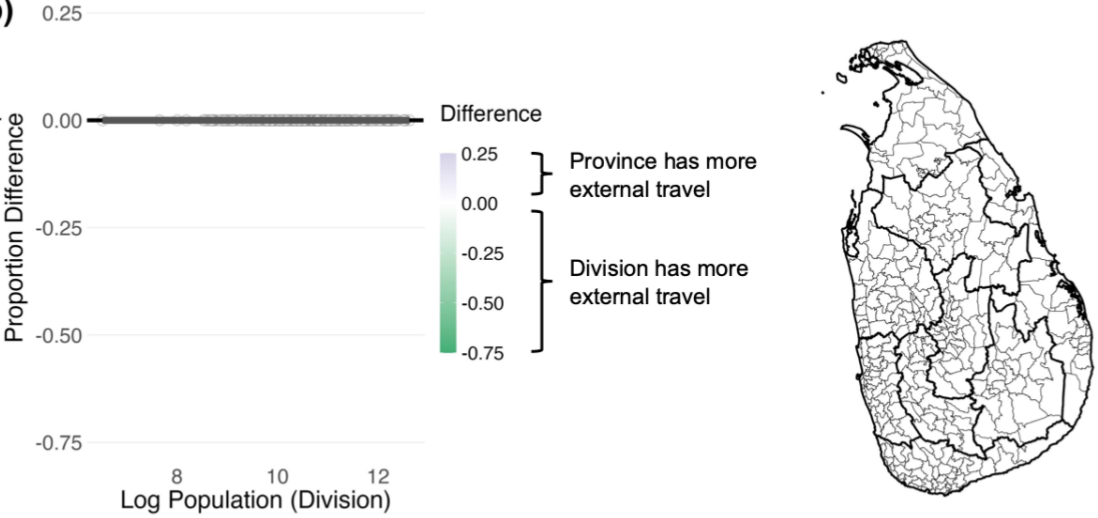


### District - Division

**(c)**

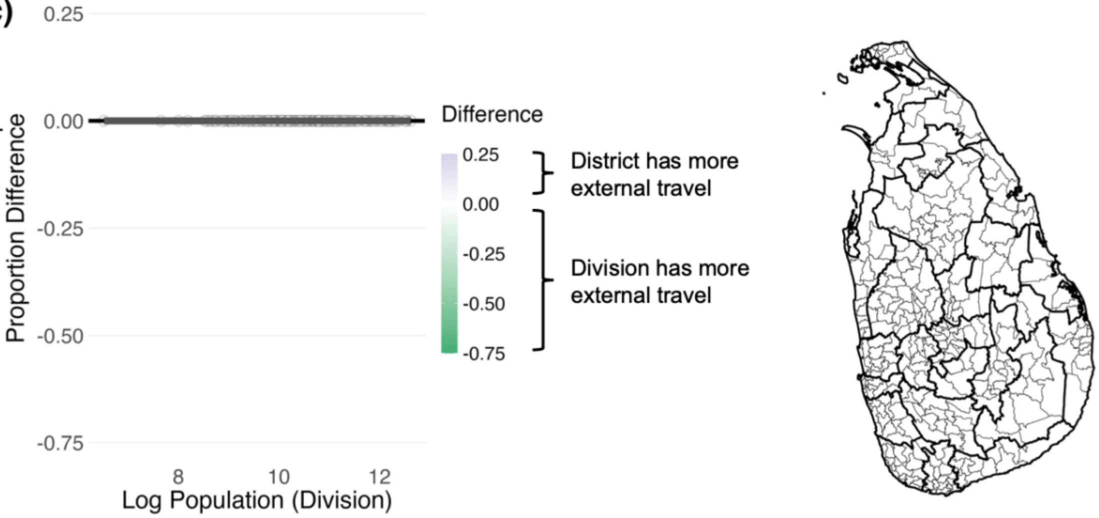

**Notes. (a)**, **(b)**, **(c)** Scatter plots with a linear trend line present the external trip proportion differences against the nested subunit population size for districts within provinces, divisions within provinces, and divisions within districts. Differences are calculated by subtracting the subunit proportion from the unit proportion. Maps display the external trip proportion differences spatially, where colors correspond to the scale that has more external travel.

### S.8 Disease Simulations at Various Spatial Scales with Observed and Rescaled Mobility Data

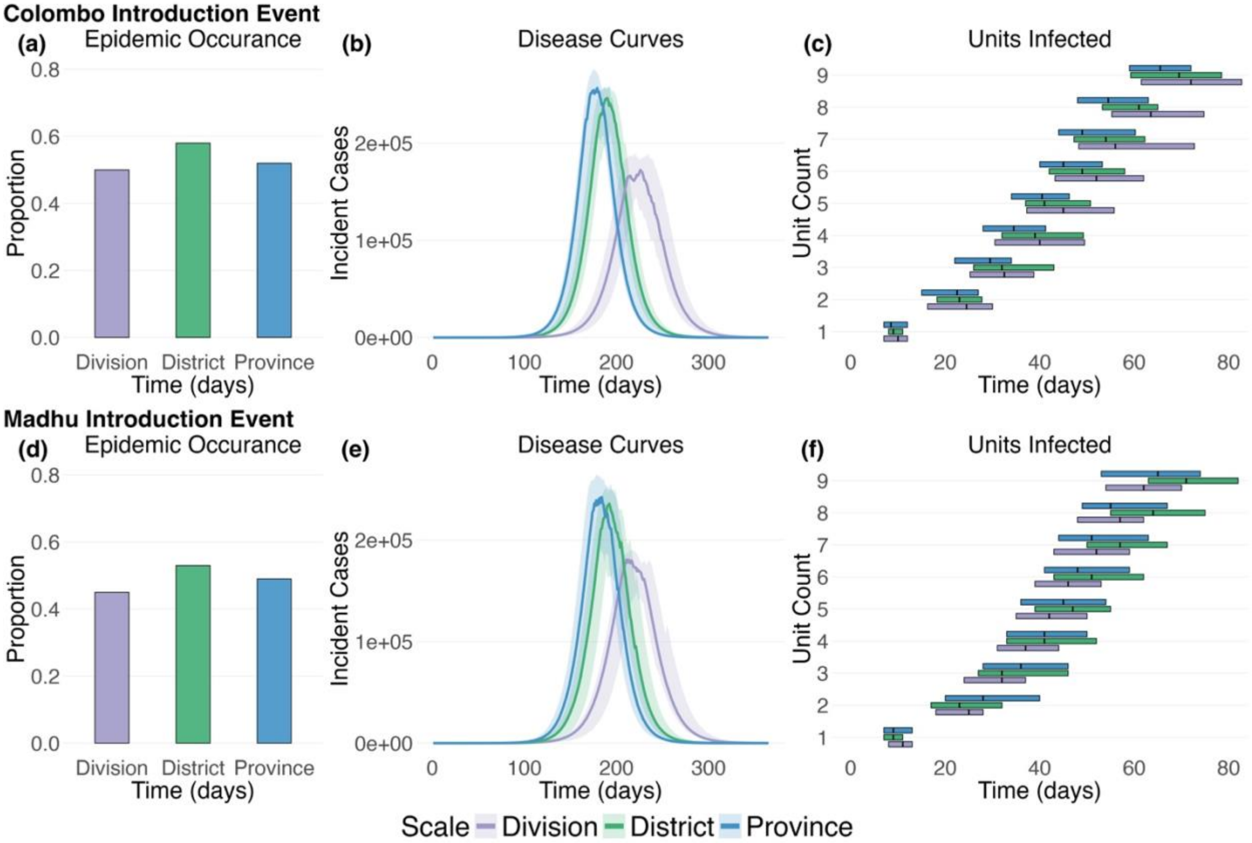


**Notes.** Plots summarize two epidemic simulation scenarios: Disease dynamics modeled with rescaled mobility considering a Colombo introduction event and disease dynamics modeled with rescaled mobility considering a Madhu introduction event. **(a), (d)** Bar plots display the proportion of disease simulations that produced more than 100 total cases (i.e. resulted in an epidemic). **(b), (e)** Epidemic curves display the 50$^{th}$, 25$^{th}$, and 75$^{th}$ percentiles of incident cases across the epidemic simulations for the entire country comparing the three different spatial scale models. **(c), (f)** Horizontal box plots show the timing and number of provinces infected as disease moves throughout the population examining the three different spatial scale models (aggregated to the province level for comparison). The 50$^{th}$, 25$^{th}$, and 75$^{th}$ percentiles for when more than 1 cumulative case occurred in each province is displayed across simulations. $R_0$ was set at 2, the latent period was set at 3 days, and the infectiousness duration was set at 5 days for each simulation.

### S.9 Pairwise Differences in Epidemic Statistics between Models Run at Various Spatial Scales with Observed and Rescaled Mobility Data

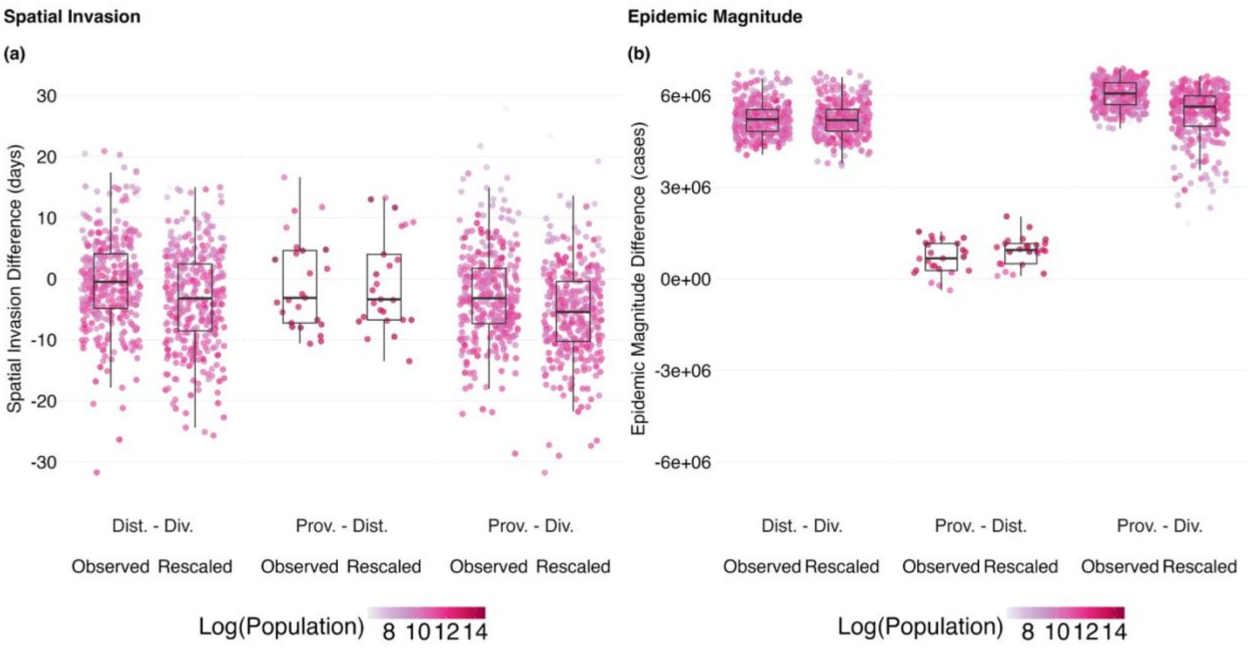


**Notes.** Each pair of boxplots compare difference distributions between observed and rescaled mobility data. **(a)** Boxplots display the differences in average spatial arrival times between models run with various spatial scales of mobility data. **(b)** Boxplots display the differences in epidemic magnitude between models run with various spatial scales of mobility data. Point colors correspond to the natural logarithm population size of the disease introduction location. Multiple pairwise spatial scale comparisons are made in each panel.

**Table S.4. Mobility Descriptive Statistics by Mobility Data Type and Spatial Scale**

| Spatial Scale | Data Type | Total Daily Average Trips | Max Daily Average Trips | Avg Trip Distance (km) | Max Trip Distance (km) |
|---|---|---|---|---|---|
| Province | Observed | 40,166,011 | 14,713,697 | 9.23 | 327.14 |
| | Simulated | 52,134,061 | 19,427,294 | 9.24 | 327.14 |
| District | Observed | 40,167,885 | 6,439,044 | 11.64 | 397.36 |
| | Simulated | 53,886,709 | 6,860,578 | 11.65 | 397.36 |
| Division | Observed | 40,987,358 | 568,713 | 17.81 | 429.79 |
| | Simulated | 40,322,976 | 130,125 | 17.94 | 429.79 |

**Notes.** Simulated data shows close agreement to observed data in terms of average daily trips, maximum daily trips, average trip distance, and maximum trip distance.

**Table S.5a. Division Discrepancies between Geographic Data and Official Sri Lankan Government Distinctions**

| Geographic Data Divisional Secretariat Name | Official Divisional Secretariat Distinction |
|---|---|
| Kalthota | Balangoda |
| Madampagama | Hikkaduwa |
| Mathurata | Hanguranketa |
| Nildandahinna | Walapane |
| Norwood | Ambagamuwa |
| Rathgama | Hikkaduwa |
| Thalawakele | Nuwara Eliya |
| Wanduramba | Baddegama |

**Table S.5b. Division Discrepancies between Mobile Phone Data and Official Sri Lankan Government Distinctions**

| Mobile Phone Data Divisional Secretariat Name | Official Divisional Secretariat Distinction |
|---|---|
| Kothmale | Kothmale East |
| Kothmale | Kothmale West |

**Notes:** Each table describes the misalignment in divisional secretariat distinctions between **(a)** geographic data and official government units and **(b)** mobile phone data and official government units. For example, in the geographic data, Hikkaduwa is divided into three units (Hikkaduwa, Madampagama, and Rathgama) while in the mobile phone data, Kothmale is a combination of two units (Kothmale East and Kothmale West). For all analyses, data sources were aligned to the 330 divisional secretariate distinctions in the mobile phone data.
